# Epidemiological and Genomic analysis of a Sydney Hospital COVID-19 Outbreak

**DOI:** 10.1101/2021.02.17.21251943

**Authors:** Elaine Tennant, Melanie Figtree, Jo Tallon, Rowena A Bull, Malinna Yeang, Ira W Deveson, James M Ferguson, Thiruni Adikari, Edward C Holmes, Sebastiaan Van Hal, Jillian M Hammond, Igor Stevanovski, Katerina Mitsakos, Drew Hilditch-Roberts, William Rawlinson, Bernard Hudson

## Abstract

Australia’s early COVID-19 experience involved clusters in northern Sydney, including hospital and aged-care facility (ACF) outbreaks. We explore transmission dynamics, drivers and outcomes of a metropolitan hospital COVID-19 outbreak that occurred in the context of established local community transmission. A retrospective cohort analysis is presented, with integration of viral genome sequencing, clinical and epidemiological data. We demonstrate using genomic epidemiology that the hospital outbreak (n=23) was linked to a concurrent outbreak at a local aged care facility, but was phylogenetically distinct from other community clusters. Thirty day survival was 50% for hospitalised patients (an elderly cohort with significant comorbidities) and 100% for staff. Staff who acquired infection were unable to attend work for a median of 26.5 days (range 14-191); an additional 140 staff were furloughed for quarantine. Transmission from index cases showed a wide dispersion (mean 3.5 persons infected for every patient case and 0.6 persons infected for every staff case). One patient, who received regular nebulised medication prior to their diagnosis being known, acted as an apparent superspreader. No secondary transmissions occurred from isolated cases or contacts who were quarantined prior to becoming infectious. This analysis elaborates the wide-ranging impacts on patients and staff of nosocomial COVID-19 transmission and highlights the utility of genomic analysis as an adjunct to traditional epidemiological investigations. Delayed case recognition resulted in nosocomial transmission but once recognised, prompt action by the outbreak management team and isolation with contact and droplet (without airborne) precautions were sufficient to prevent transmission within this cohort. Our findings support current PPE recommendations in Australia but demonstrate the risk of administering nebulised medications when COVID-19 is circulating locally.

## Introduction

The COVID-19 pandemic is a major threat to human physical and mental health. A relatively short incubation period, pre-symptomatic viral shedding and immunologically naïve population all facilitate rapid propagation(1, 2). A proportion of cases require tertiary care, with quoted hospitalisation rates between 0.8 and 20% globally(3, 4). Those not needing admission may still attend healthcare facilities for assessment. Hospitals therefore represent high risk areas for transmission.Healthcare workers (HCWs) can suffer illness and additionally become vectors, unwittingly endangering vulnerable patients.

Australia identified its first COVID-19 cases in late January after which a travel ban to China was implemented. Roughly one month later further cases occurred, including within Northern Sydney Local Health District (NSLHD). Here, during March/early April, a hospital outbreak and several community clusters emerged, including Australia’s first major residential aged-care facility (RACF) outbreak (5). Nosocomial transmission soon became evident within the hospital. Herein, we perform an analysis of the hospital outbreak, merging the findings of contact tracing operations with SARS-CoV-2 genomic epidemiology.

## Methods

### Outbreak setting and routine control measures

The outbreak occurred at Ryde Hospital, a 174 bed general facility situated in metropolitan Sydney, for which our team provide infectious diseases (ID)/microbiology consultation services. The first patient met confirmed case definition for COVID-19 infection on 2^nd^ March 2020 and the last on 2^nd^ April 2020. On 25^th^ February, when the first case was suspected, an outbreak management team (OMT) was formed, comprising Infection Prevention and Control Practitioners (IPCP), senior departmental staff and the consulting ID specialist team. Support was provided by NSLHD Public Health Unit(PHU) and hospital executive.

Case management was informed by the Communicable Diseases Network Australia Series of National Guidelines (CDNA SONG) on management of COVID-19(6). These continue to be revised regularly; the most recent iteration was referenced at all times. Confirmed cases had a positive molecular test for SARS-CoV-2 RNA on nasopharyngeal swab, sputum or broncho-alveolar lavage. Initial testing was performed at SAViD (Serology and Virology Division NSW Health Pathology) and subsequently at Royal North Shore Hospital once in-house testing capability was established. Real-time PCR detected molecular targets including E gene, RdRp, N gene and/or Orf 1b. SARS-CoV-2 genome sequencing was attempted routinely on all positive samples.

All suspected and confirmed cases were managed with combined contact, droplet and standard precautions as per NSW Clinical Excellence Commission (CEC) guidelines(7). These comprised isolation in a single room and staff PPE comprising surgical mask, fluid-resistant long-sleeved gown, gloves and eye protection. Airborne precautions were implemented if an aerosol generating procedure was required. Once nosocomial transmission was recognised, active case detection was undertaken for all staff (by questioning for symptoms at the start of every shift) and patients (by identifying patients with “hospital acquired pneumonia” on the electronic Antimicrobial Stewardship sYstem (eASY AMS; Monitor Software, Sydney, Australia).

At the time, all COVID-19 cases diagnosed within NSLHD were notified to the ID/Clinical Microbiology team for management and staff residing in other LHDs identified as cases/contacts were notified to the OMT via district infection control. Hospital-based patient contacts were identified using a dedicated contact tracing application (QlikQ, QlikTech International AB) that integrated with the electronic medical records, and by direct communication with health managers. Given the high risk clinical setting, staff identified as contacts were instructed to self-isolate for 14 days and present for testing should they develop symptoms. Clearance/release from isolation was granted per CDNA Guidelines.

### Retrospective outbreak analysis

A retrospective analysis of pre-collected data was undertaken, with three aims: first, to define the hospital associated outbreak cohort; second, to clarify any links between the hospital outbreak and concomitant community clusters; third, to examine the drivers of hospital associated transmission. Research approval was granted by NSLHD Human Research Ethics Committee (HREC) (2020/STE01571). An epidemiologic outbreak hypothesis and viral phylogenetic trees analysis were developed independently to avoid bias. The clinical and molecular teams then met to refine the outbreak hypothesis.

### Definition of the outbreak cohort and locally acquired clusters (Fig 1)

The OMT was aware of 63 persons diagnosed with COVID-19 between 2^nd^ March and 2^nd^ April 2020 within the hospital and/or surrounding community. Their clinical records were screened to determine likely source of acquisition and seek any hospital contact during their incubation/infectious periods. Hospital attendance for COVID-19 testing (performed under contact and droplet precautions) was not included. 49 persons were determined to have locally-acquired transmission. Of these, 23 were implicated in the hospital transmission and/or acquisition and were included in a focused outbreak analysis.

**Fig 1:**
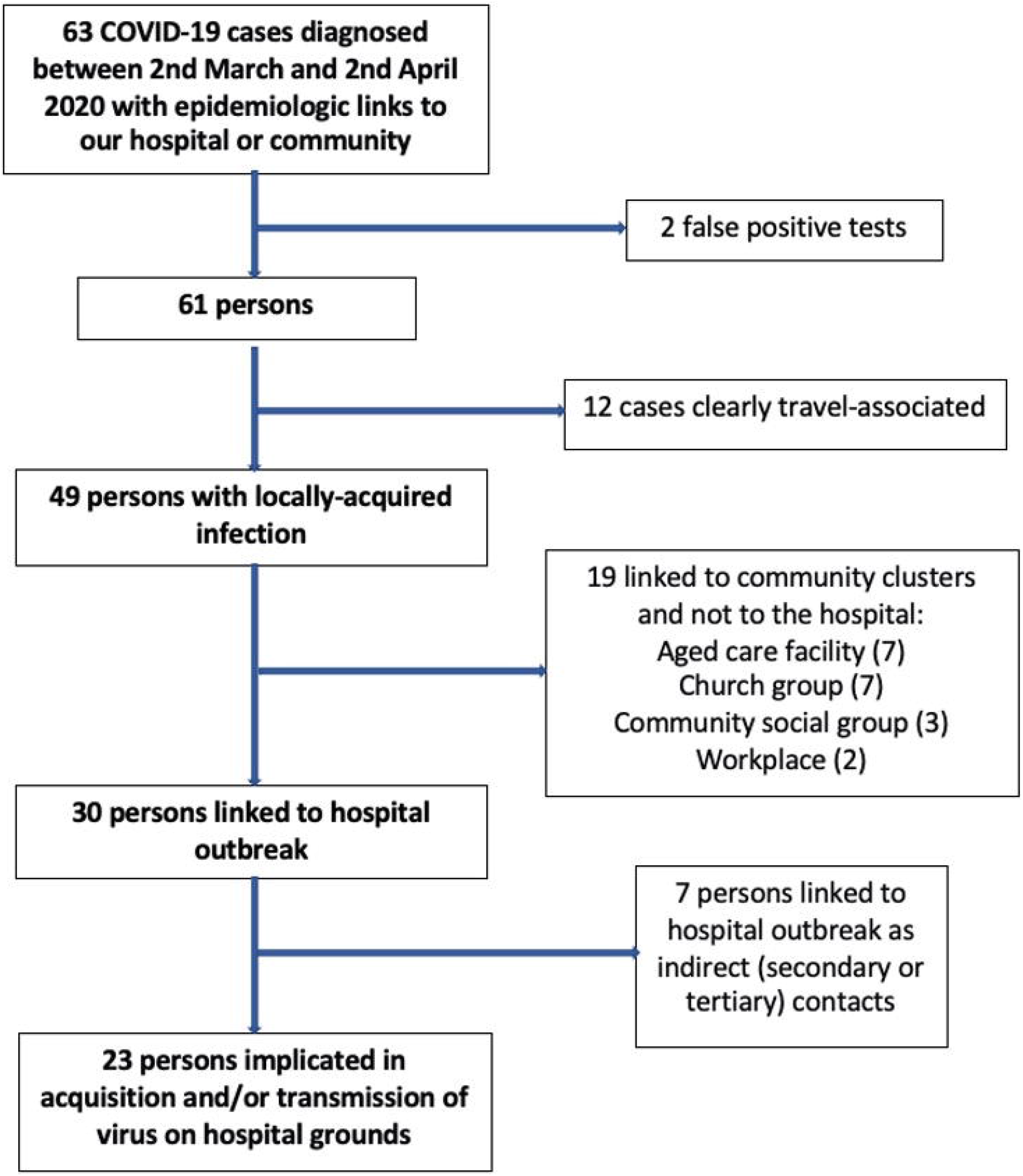
Flow diagram for defining a cohort involved in a hospital-associated outbreak of COVID-19, Sydney, 2020.

### SARS-CoV-2 genomic epidemiology

We sought to obtain SARS-CoV-2 genome sequences for all persons implicated with locally acquired infection (n=49 samples of interest). Accordingly, SARS-CoV-2 genome sequence data was obtained for all but one of the samples and performed by UNSW SAViD (Serology and Virology Division NSW Health Pathology) team, according to an analytically validated workflow employing Oxford Nanopore sequencing (8). The methods are described in the Supporting information section. One patient who was transferred to Westmead Hospital had genome sequencing performed on their sample by NSW Health Pathology (NSWHP) at that site.

### Hospital outbreak analysis

A detailed analysis was undertaken of the 23 persons involved in the hospital outbreak. Information collected included age, comorbidities, date of symptom onset, implementation of contact/droplet precautions, tests undertaken(positive and negative) and clinical outcome. Information was gathered about movement of each person within clinical areas and period of isolation/absence from work.

For all identified cases, exposure periods (based on an incubation period of 1-14 days) and infectious periods (beginning 48 hours prior to symptom onset until date of certified clearance) were determined(6). Likely hospital transmission routes were hypothesised using this information. Number of secondary transmissions were calculated based on maximum number of plausible transmissions from a single person.

## Results

### Cohort Identification: Epidemiologic and Genomic Analysis

Of the 23 hospital outbreak cases, 13 samples underwent successful viral genome sequencing and were identified as belonging to SARS-CoV-2 lineage B4 using the Pangolin lineage assignment tool (Fig 2). Additionally, in two cases there was a clear epidemiological link between index cases and secondary/tertiary generations from whom genome sequence data was obtained: the lineage of the index person was also inferred as B4. Overall, viral genome sequence data was determined in 9/11 patients/visitors and 7/12 staff. Three of the patients were residents of the local ACF involved in the outbreak and their viral genome sequences clustered with other hospital cases (also within lineage B4).

**Fig 2:**
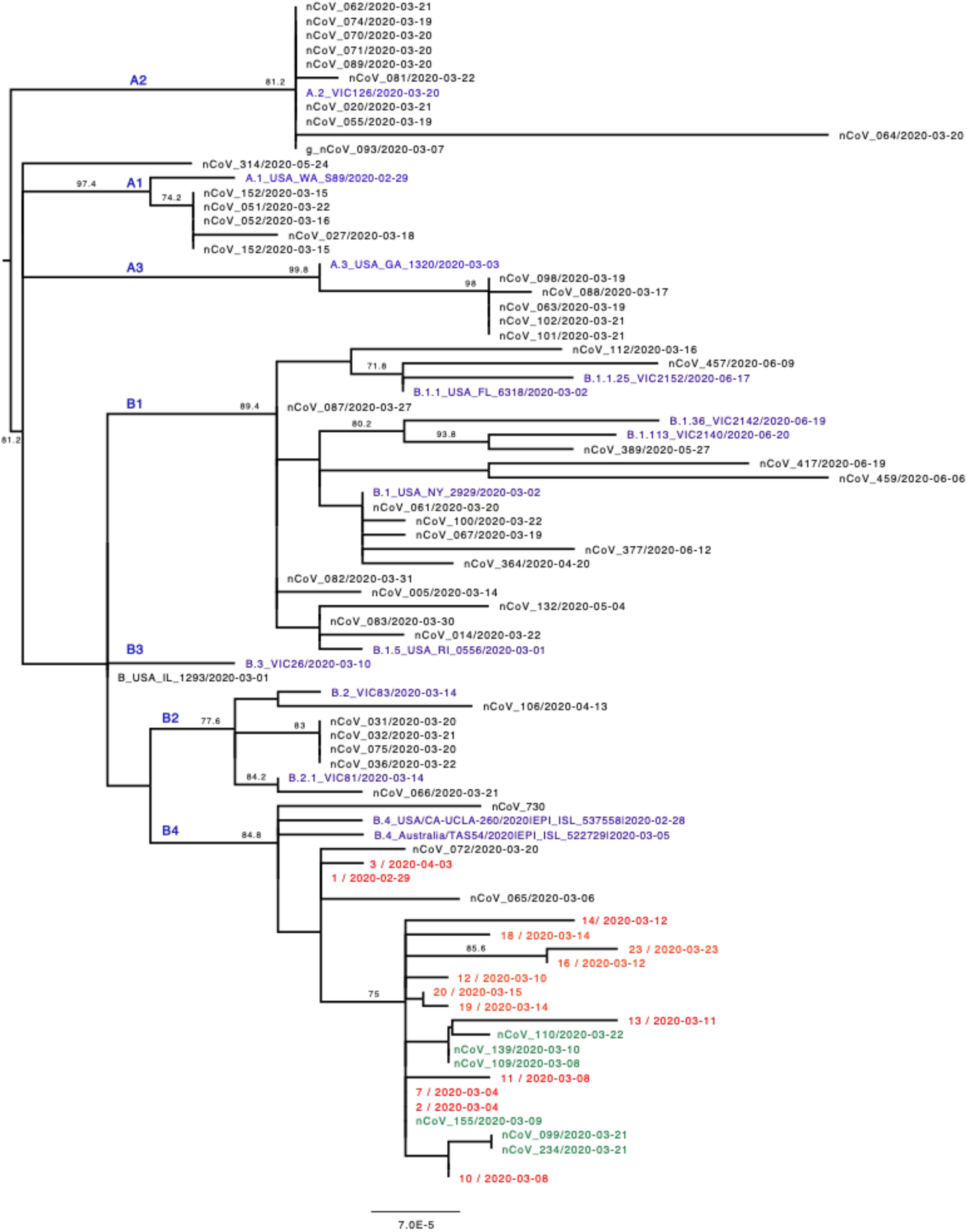
Phylogenetic analysis of an outbreak of COVID-19 at Ryde hospital, Sydney during March-April 2020. Red: Hospital outbreak associated cases labelled by their identification number followed by the date of sample isolation; Green: Cases from local RACF outbreak but not admitted to hospital; Blue: Reference strains denoting lineages from GISAID; Black: Other SARS-CoV-2 strains circulating in NSW at the time. The percent bootstrap values in which the major groups were observed among 1000 replicates are indicated. The phylogenetic tree was midpoint rooted for clarity and the scale bar denotes the number of nucleotide substitutions per site.

A further 26 persons had locally-acquired infection during the outbreak period. Seven of these were indirectly linked to the hospital (through household/social contact), 5 of which were fell within the B4 lineage. A further 7 were linked to the ACF but not the hospital; 3 these were sequenced and all found to be lineage B4. In contrast, 8/12 samples from other concurrent community outbreaks in NSLHD were sequenced; these were phylogenetically distinct from the ACF/hospital cases, falling into the B1 and A3 lineages.

### Hospital cases: Clinical characteristics and outcomes

The hospital outbreak cohort included 12 staff members, 10 patients (8 of whom were hospitalised with COVID-19 infection) and one visitor to a later diagnosed COVID-19 patient. One patient was also a staff member. Fig 3 shows the epidemic curve of the outbreak and Fig 4 shows the outbreak timeline with exposure and infectious periods. Table 1 summarises the characteristics of infected patients/visitors and staff, while clinical, virologic and transmission characteristics of each infected person are presented in Tables 2 and 3. Patients/visitors affected were advanced in age (median 81 years, range 54-91) compared to infected HCWs (median 42 years, range 19-66) and had more comorbid conditions.

**Table 1:** Comparison between hospitalised patients and infected staff amongst a group implicated in a hospital-associated COVID-19 outbreak in a Sydney Hospital, 2020.

|  | Patients/Visitors* | Hospital Staff |
| --- | --- | --- |
| **Patient Characteristics and Outcomes:** |  |  |
| Age, median (range) | 81 (54-94) | 42 (19-66) |
| Occupation (for affected staff only) | N/A | Registered nurse (4)<br>Assistant in nursing (1)<br>Doctor (4)<br>Occupational therapist (1)<br>Physiotherapist (1)<br>Food services (1)<br>Cleaner (1) |
| Medical comorbidities | Thromboembolic (1/11)<br>Malignancy (1/11)<br>Obstructive airways disease (2/11)<br>Hypertension (3/11)<br>Haemochromatosis (2/11)<br>Ischaemic heart disease (1/11)<br>Cardiac arrhythmia (2/11)<br>CCF (1/11)<br>Vasculopathy (IHD/PVD/CVA) (2/11)<br>Other Lung disease (4/11)<br>Hypercholesterolaemia (3/11)<br>Overweight (2/11) | Thromboembolic (1/12)<br>Malignancy (1/12)<br>Obstructive airways disease (1/12)<br>None (10/12) |
| Hospitalisation rate | 72.7% (8/11) | (0%) 0/12 |
| 30 day survival rate of infected patients (%) | 63.6%<br>(7/11) | 100%<br>(12/12) |
| 30 day survival rate amongst hospitalised patients (%) | 50.0%<br>(4/8) | N/A |
| Time lost from work in days; median (range) | N/A | 26.5<br>(14-191) |
| **Transmission Characteristics** |  |  |
| Incubation period (days) | Unknown: 7/11 (70%)<br>Median: 6.6<br>Mean: 5.8<br>Range: 4-7 | 3/12 (25%)<br>5.0<br>5.7<br>2-11 |
| Number of days spent on hospital grounds during infectious period, median (range) | 1 (0-7) | 0 (0-5) |
| Mean number of secondary transmissions (range): | 3.5<br>(0-11) | 0.6<br>(0-3) |

\* Patient who was also a staff member included within patient/visitor group for all categories, other than "Number of days spent on hospital grounds during infectious period, which is separated out as days spent as staff and days spent as patient in appropriate categories). This person is also included amongst staff in the "Time lost from work" category.

**Table 2:**
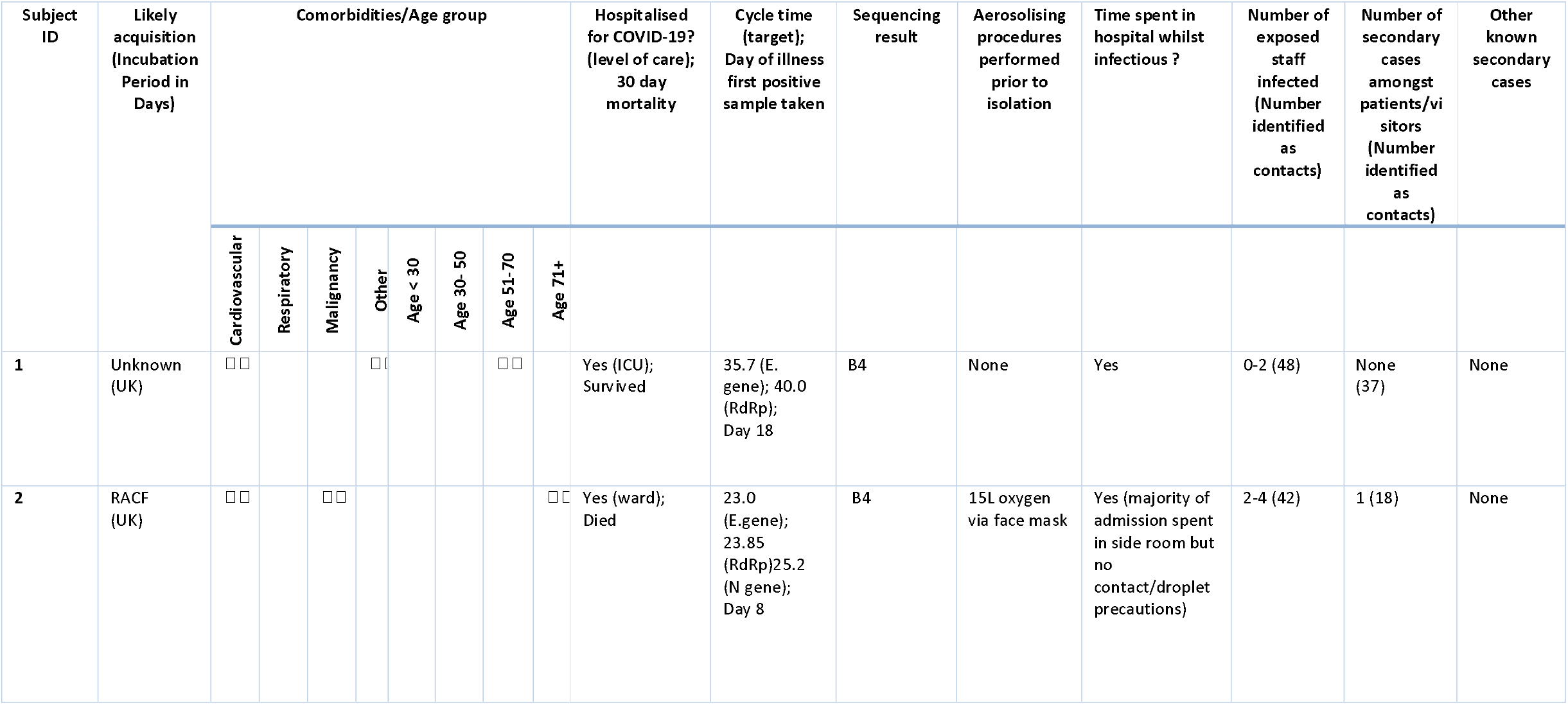
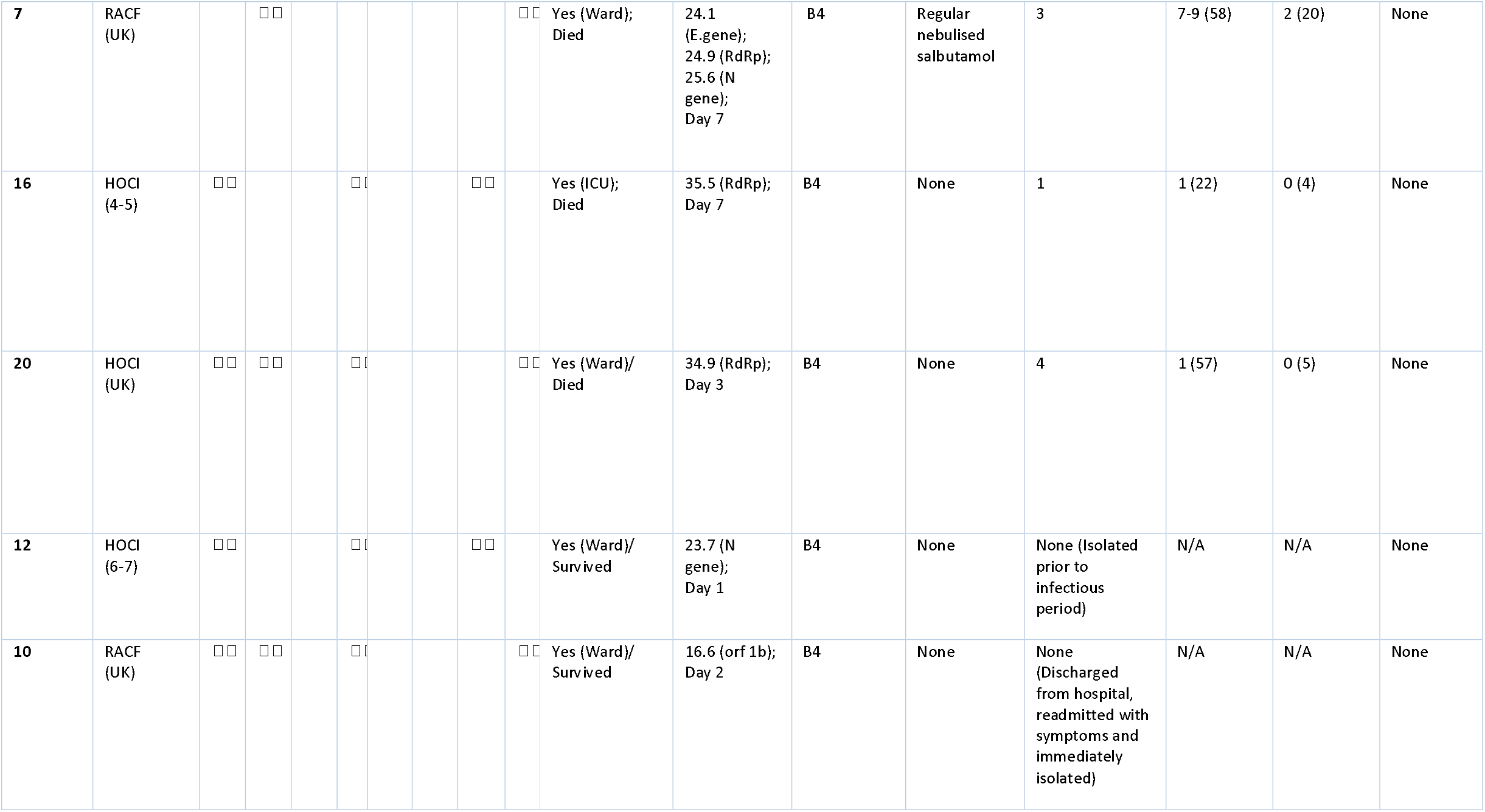
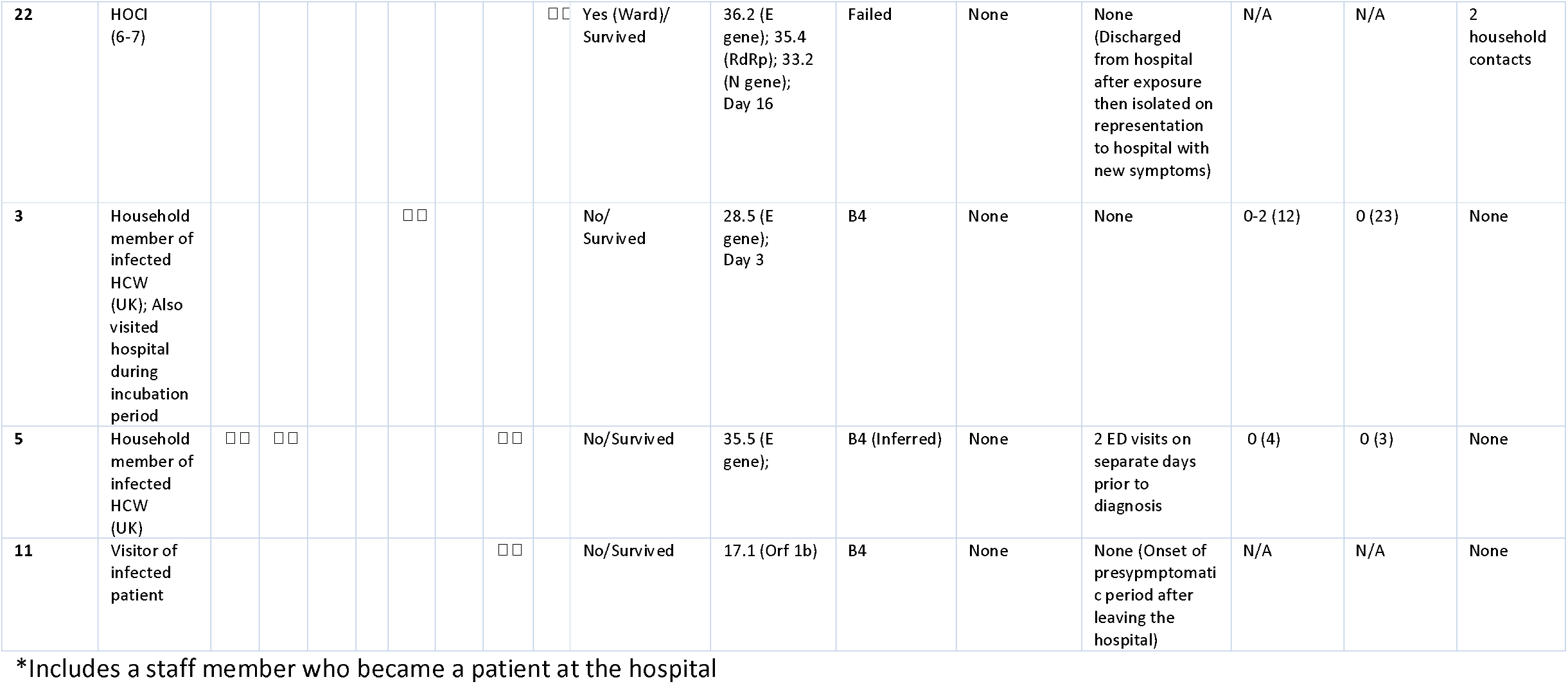
Summary of patients/visitors involved in an outbreak of COVID-19 at a Sydney hospital, March-April 2020. UK = Unknown

**Table 3:**
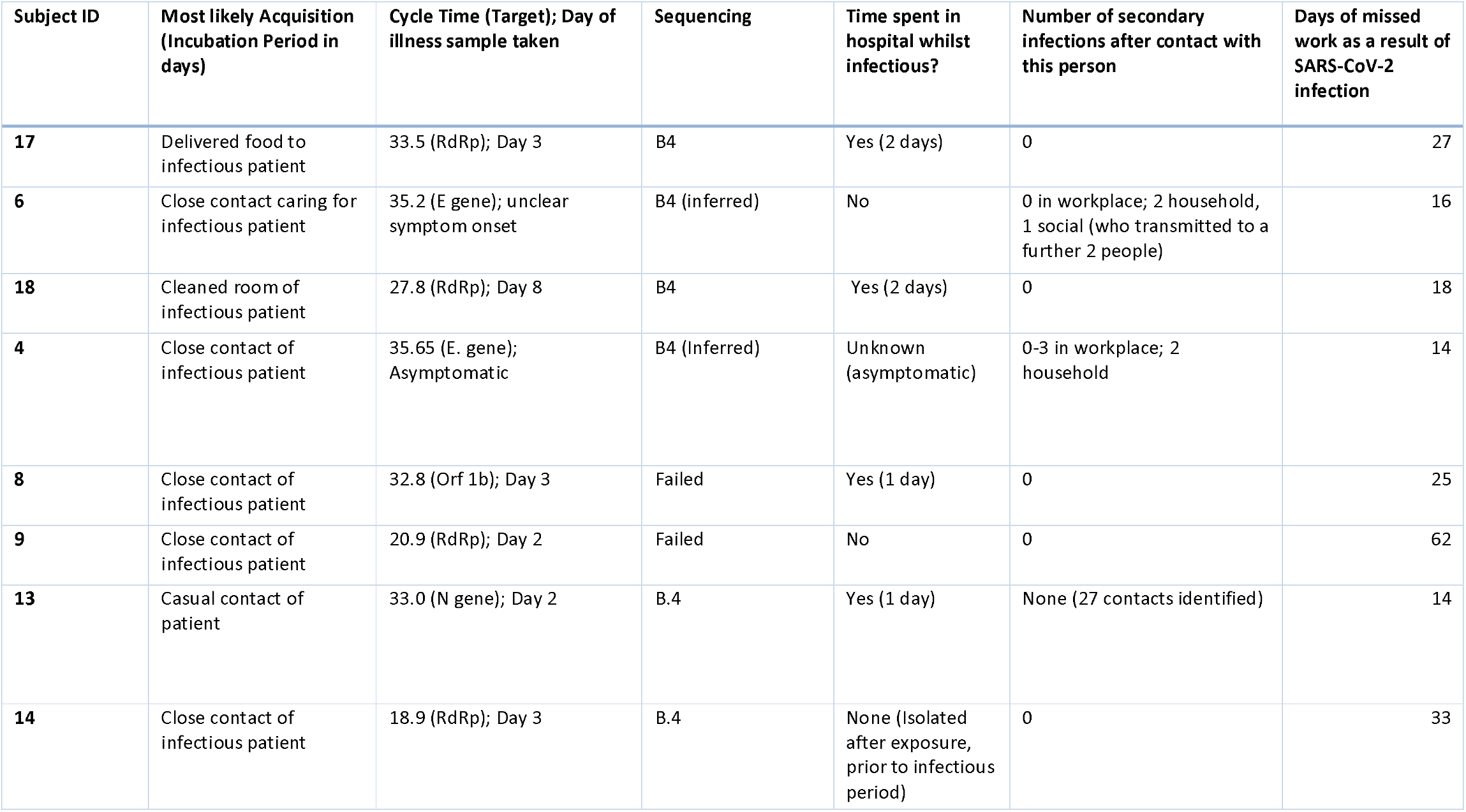
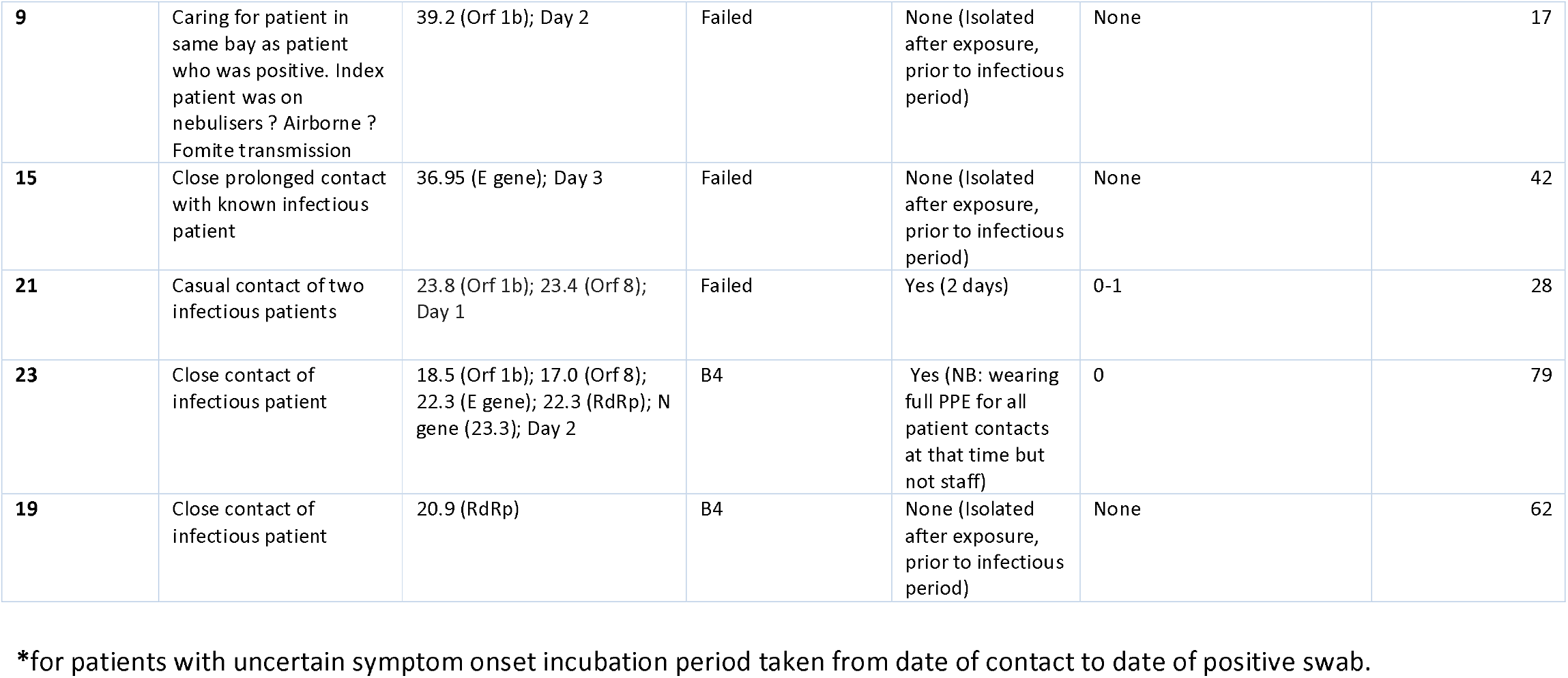
Summary of staff members involved in an outbreak of COVID-19 at a Sydney hospital, March-April 2020.

| Subject ID | Most likely Acquisition (Incubation Period in days) | Cycle Time (Target); Day of illness sample taken | Sequencing | Time spent in hospital whilst infectious? | Number of secondary infections after contact with this person | Days of missed work as a result of SARS-CoV-2 infection |
| --- | --- | --- | --- | --- | --- | --- |
| 17 | Delivered food to infectious patient | 33.5 (RdRp); Day 3 | B4 | Yes (2 days) | 0 | 27 |
| 6 | Close contact caring for infectious patient | 35.2 (E gene); unclear symptom onset | B4 (inferred) | No | 0 in workplace; 2 household, 1 social (who transmitted to a further 2 people) | 16 |
| 18 | Cleaned room of infectious patient | 27.8 (RdRp); Day 8 | B4 | Yes (2 days) | 0 | 18 |
| 4 | Close contact of infectious patient | 35.65 (E. gene); Asymptomatic | B4 (Inferred) | Unknown (asymptomatic) | 0-3 in workplace; 2 household | 14 |
| 8 | Close contact of infectious patient | 32.8 (Orf 1b); Day 3 | Failed | Yes (1 day) | 0 | 25 |
| 9 | Close contact of infectious patient | 20.9 (RdRp); Day 2 | Failed | No | 0 | 62 |
| 13 | Casual contact of patient | 33.0 (N gene); Day 2 | B.4 | Yes (1 day) | None (27 contacts identified) | 14 |
| 14 | Close contact of infectious patient | 18.9 (RdRp); Day 3 | B.4 | None (Isolated after exposure, prior to infectious period) | 0 | 33 |
| **9** | Caring for patient in same bay as patient who was positive. Index patient was on nebulisers ? Airborne ? Fomite transmission | 39.2 (Orf 1b); Day 2 | Failed | None (Isolated after exposure, prior to infectious period) | None | 17 |
| **15** | Close prolonged contact with known infectious patient | 36.95 (E gene); Day 3 | Failed | None (Isolated after exposure, prior to infectious period) | None | 42 |
| **21** | Casual contact of two infectious patients | 23.8 (Orf 1b); 23.4 (Orf 8); Day 1 | Failed | Yes (2 days) | 0-1 | 28 |
| **23** | Close contact of infectious patient | 18.5 (Orf 1b); 17.0 (Orf 8); 22.3 (E gene); 22.3 (RdRp); N gene (23.3); Day 2 | B4 | Yes (NB: wearing full PPE for all patient contacts at that time but not staff) | 0 | 79 |
| **19** | Close contact of infectious patient | 20.9 (RdRp) | B4 | None (Isolated after exposure, prior to infectious period) | None | 62 |

\*for patients with uncertain symptom onset incubation period taken from date of contact to date of positive swab.

**Fig 3:**
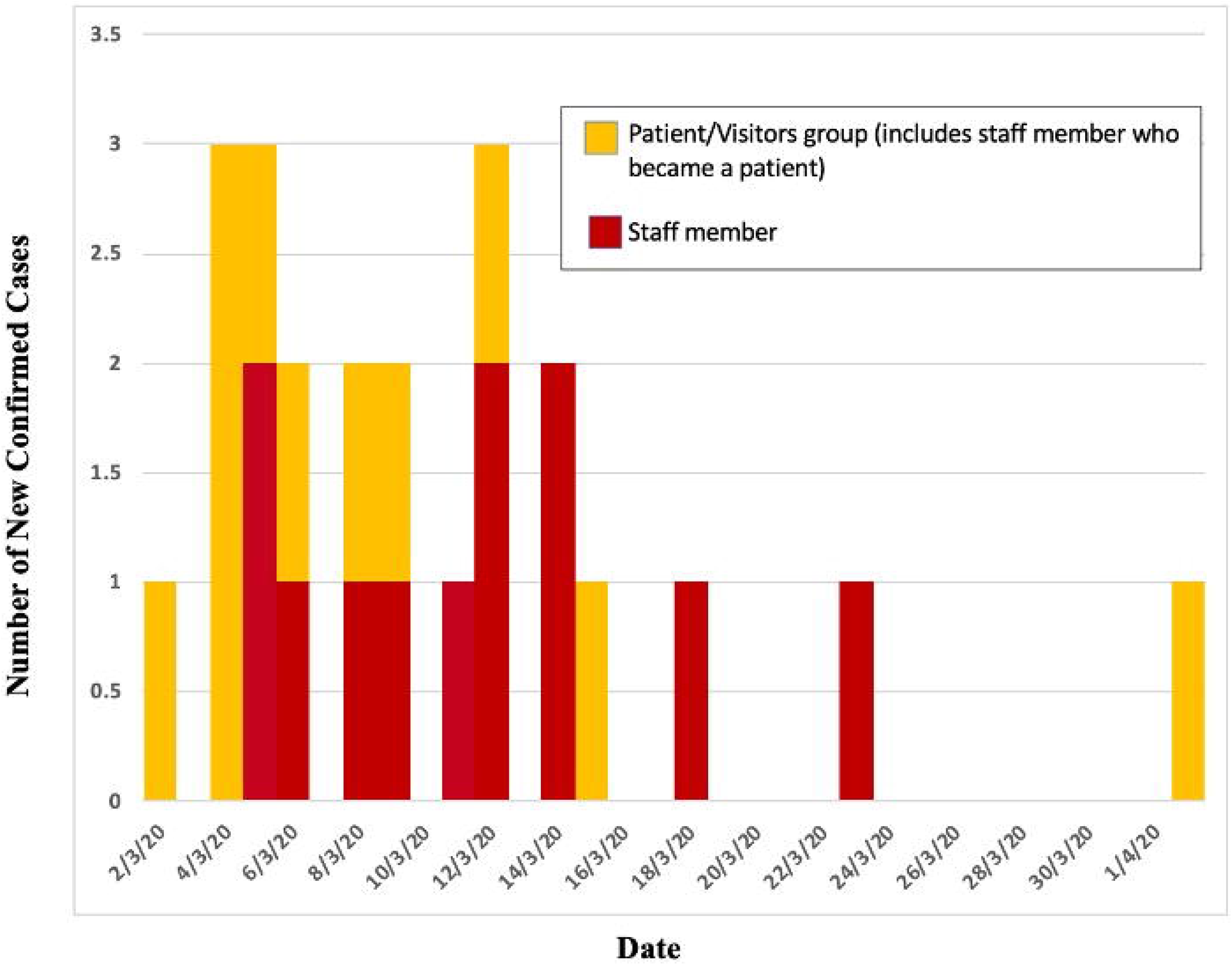
Epidemic curve for Hospital associated COVID-19 outbreak in a Sydney Hospital, March-April 2020.

**Fig 4:**
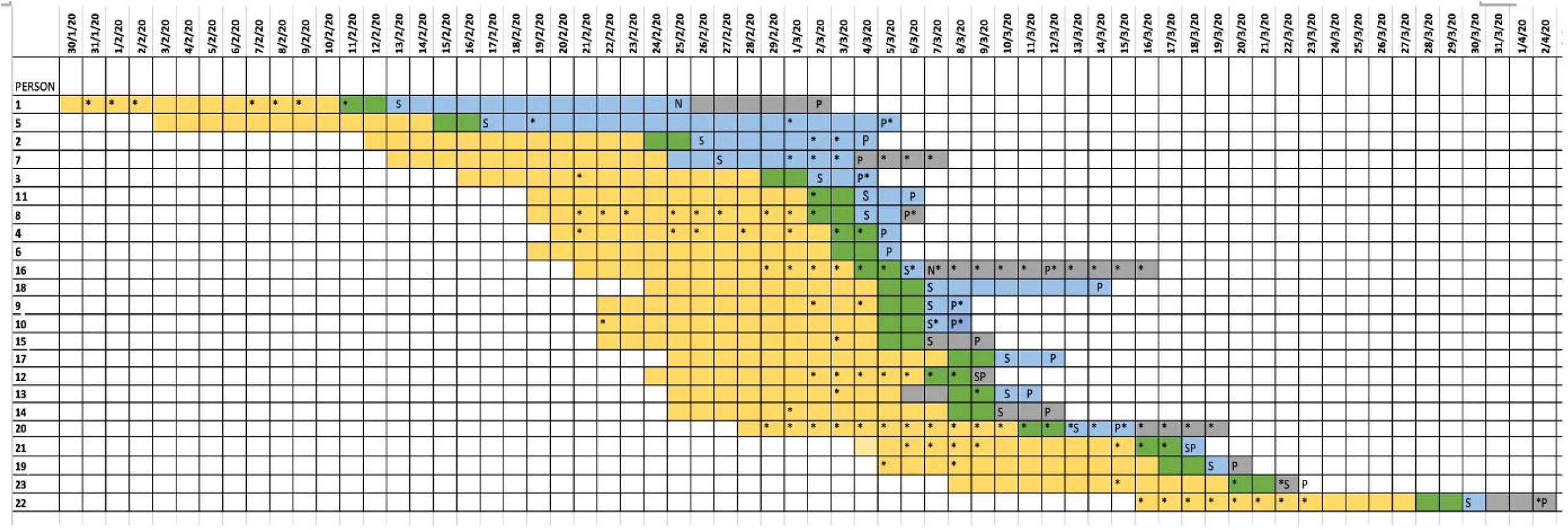
Timeline of cases in a hospital associated outbreak of COVID-19, Sydney March-April 2020. X-axis: Date; Y axis: Infected individual S : Onset of symptoms; P: Positive COVID-19 test; N: Negative COVID-19 test; *indicates person was present on hospital grounds that day. Shading: Yellow-Potential period of exposure (person within their incubation period); Green – Potential exposure or presymptomatic infectious period; Blue: Infectious period (person not on contact precautions/isolation); Grey: Infectious period (Person on hospital grounds but contact precautions implemented)

Thirty day survival was 63.6% amongst the patients/visitors groups and 50.0% amongst those who were hospitalised with COVID-19. All infected HCWs survived at 30 days, although one was hospitalised, requiring ICU level care. There was substantial forced absenteeism amongst infected staff (median 26.5 days, range 14-191 days). Additionally, a total of 140 staff (71 nurses, 31 doctors, 15 porters, 12 cleaners, 10 allied health professionals and one administration officer) were identified as contacts and furloughed for quarantine for between one and 18 days.

### Hospital transmission characteristics

The median incubation period was 6.6 days for patients and 5.0 days for staff (range 2-11 days) (Fig 5). Several patients/staff unwittingly spent time on site whilst infectious. One patient was administered regular nebulised medication in a four bedded room for several days prior to their diagnosis being known. This person’s infection resulted in the greatest number of secondary transmissions (eight). Among those infectious whilst on-site, several potential contacts were identified (range 4-58 people). The mean number of secondary cases was 3.5 for patients and 0.6 for staff, with a wide dispersion in the number of secondary transmissions (Fig 6). Among those persons identified as contacts and isolated, no secondary cases occurred. All identified transmission events could be explained by a person either coming into direct contact with an infected person without transmission-based precautions, or due to a documented PPE breach. In the remaining case, a HCW was treating a patient in an adjacent bed to the infectious person using a nebuliser.

**Fig 5:**
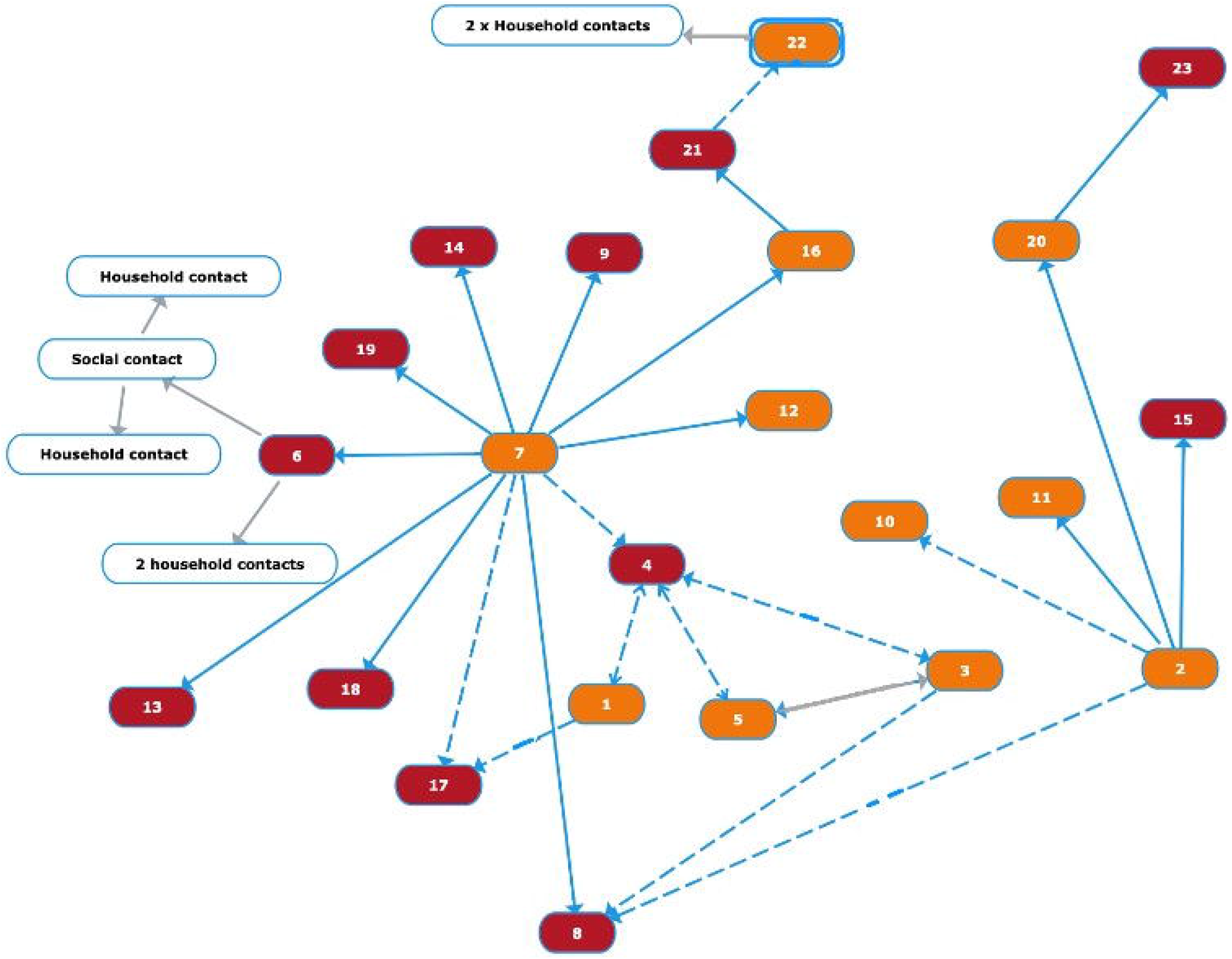
Hypothesised transmission routes between patients, staff and visitors implicated in a hospital associated COVID-19 outbreak, 2020, based on known contacts between persons during their infectious and/or incubation periods. Key: Red box: Staff member; Orange box: Person who spent time in the hospital as a patient or visitor (includes a staff member who became a patient); Clear box: Second/third generation cases as a result of this outbreak. Dashed blue line: Possible route of transmission; bold blue line: Clear route of transmission, grey line: transmission occurred out of hospital grounds

**Fig 6:**
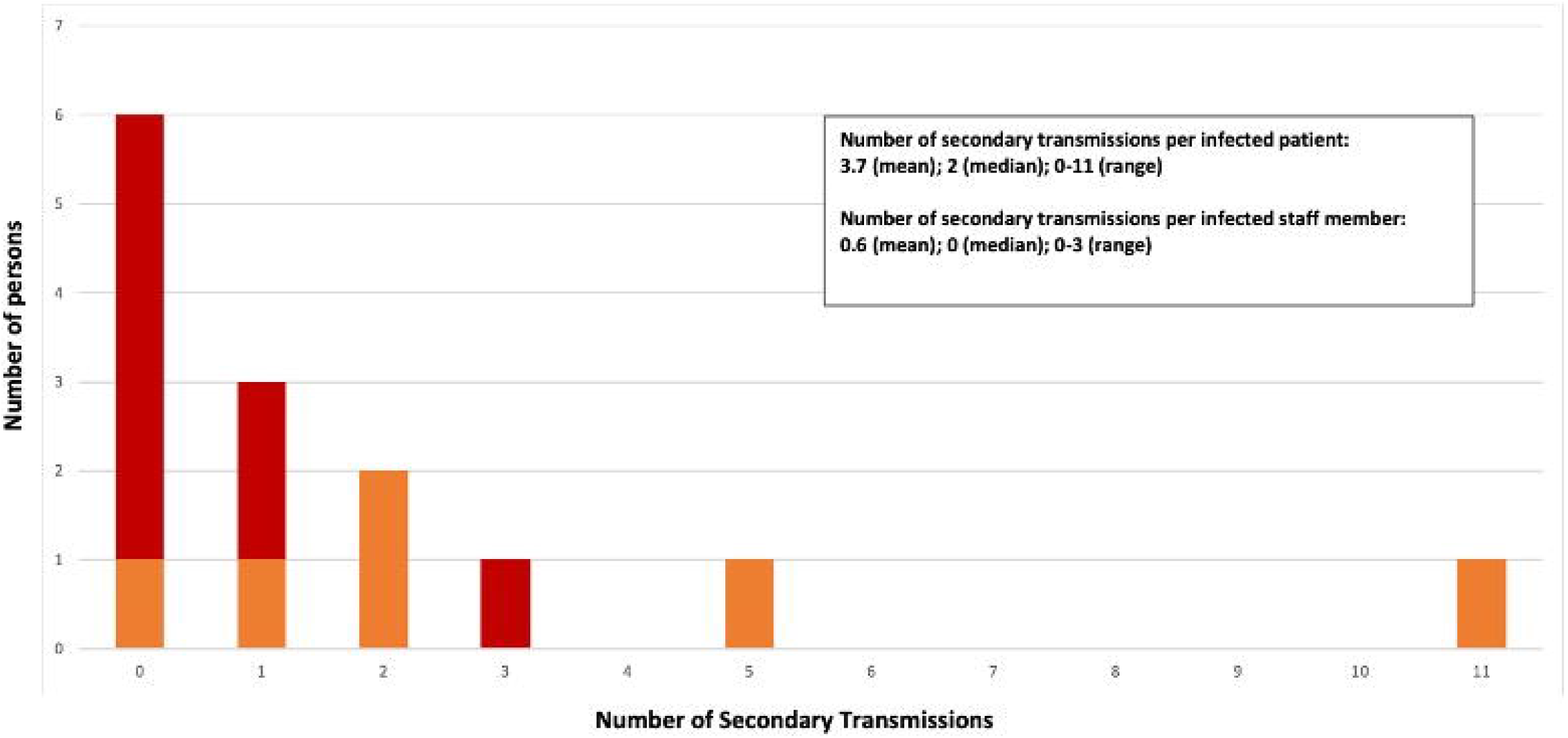
Number of secondary transmissions generated by each staff member (red) and patient (orange) infected within a hospital associated COVID-19 outbreak, Sydney 2020.

## Discussion

We present a detailed case study of a situation that played out globally in healthcare facilities throughout 2020. This outbreak was the first recognised Australian co-cluster between an ACF and a hospital. Our study provides important insights, by integrating viral genome sequence data with metadata collected by the locally-based OMT. Genomic epidemiology is an important supplement to outbreak investigations, particularly for SARS-CoV-2, and given the emergence of different viral lineages (9, 10). Although the relatively low evolutionary rate of SARS-CoV-2 usually precludes a detailed analysis of person-person transmission within outbreaks, because virus transmission occurs more rapidly than virus evolution, it does show utility in clarifying linked cases, as we have demonstrated here.

The outbreak described here began early in the Australian COVID-19 experience and some of our patients were not recognised as being infected for several days. This undoubtedly resulted in transmission episodes that were avoidable. The fact that COVID-19 symptoms overlap with other respiratory illnesses and that elderly patients can be asymptomatic/display atypical symptoms added to delay in diagnosis at this time (5)

Our key findings are threefold. First, we demonstrate the impact of hospital onset SARS-CoV-2 infection (HOCI) on patients and hospital staff. Second, we show that secondary attack rates were fairly low in this cohort (despite large numbers of contacts), yet highly variable. Finally, we describe some of the factors potentially associated with transmission and outbreak outcomes.

Nosocomial transmission of SARS-CoV-2 is of great concern to patients and HCWs(11). However, when infection is diagnosed in a HCW, hospital-based acquisition cannot always be assumed, particularly when community transmission rates are high (12). Viral genome sequencing of our outbreak cohort was useful in providing supportive evidence for our hypothesis that the identified hospital-associated cases were linked, both to each other and the local ACF outbreak. The consequences of this outbreak for individuals were significant, and in some instances devastating. Two patients who acquired the infection whilst admitted for another condition died with COVID-19. Several infected staff required sustained periods of absence from work (due to illness or prolonged SARS-CoV-2 positivity) and even more were quarantined following exposure. Although understandable from a public health perspective, prolonged absences can result in personal, financial and psychological stress to those affected. The pressure on remaining staff can increase the risk of mistakes (especially in correct use of PPE) and might encourage presenteeism, both of which may be counterproductive(13, 14). Outbreak management must therefore be finely tuned, to avoid propagation of the infection, whilst keeping the hospital running safely. When this outbreak occurred, infected staff were required to have two consecutive negative SARS-CoV-2 swabs to return to work, but with emerging evidence, these criteria have since become less stringent(6, 15).

Our study highlights the numerous social contacts that occur within hospitals, as evidenced by the numbers of exposed persons found on contact tracing. Despite this, most identified contacts did not subsequently develop COVID-19. It was notable that the number of secondary transmissions generated by each case were highly variable (between 0 and 12) and were different for patients and staff (mean 0.58 from staff; 3.25 from patients). We also observed clear overdispersion in the patterns of transmission, in which most infected persons infect no other individual, whereas as a small number of other persons act as apparent “superspreaders”. This phenomenon characterises many infections and seems particularly relevant to COVID-19 (1, 16–18).

The patient who transmitted to most others was administered medication via a nebuliser whilst infectious, an aerosol generating procedure which likely increases transmission risk (19). In healthcare settings, when there is an epidemiologic risk that COVID-19 is circulating, we would argue that nebulisers should only be used under airborne precautions and that alternatives (such as bronchodilators delivered via spacer) are utilised. There may have been other factors that facilitated spread, such as high viral shedding (not measured) and the time in hospital prior to isolation (three days). One secondary case that arose from this person was a HCW without direct contact who was treating a patient in the same bay. This may have been due to airborne or fomite transmission, although the latter likely occurs at relatively low frequency (20, 21). With the exception of the nebuliser-associated transmission (for which airborne precautions would have been used had the infection been recognised), we did not demonstrate transmission after implementation of appropriate contact and droplet precautions. This provides some reassurance regarding the current advice for use of surgical masks rather than respirators(14).

Several factors may have influenced the outcome of this outbreak. The cohort studied included patients with poor prognostic factors, including advanced age and cardiovascular/respiratory comorbidities(22). We have illustrated that a brisk outbreak management team response is critical. As well as isolating known cases, identification and quarantine of contacts prior to their infectious period is essential, given the known viral shedding profiles and pre-symptomatic transmission (23, 24). This is highlighted by the absence of secondary transmissions amongst those of our contacts who were identified and put on contact/droplet precautions/removed from work prior to becoming unwell.

Our study has several limitations. The OMT were required to gather epidemiological data rapidly. Accordingly, there may have been gaps in the collected information, due to ascertainment and recall bias. Review of the outbreak management underlines the siloed nature of our healthcare system, with allied professions being managed through very separate streams; this can present a barrier to rapid infection control actions. An OMT should incorporate cross-discipline membership to produce equivalent contact tracing and communication efforts across medical, nursing, allied health and corporate services. A further challenge is the rapid bed movement of patients and the lack of a clear formal mechanism for identifying contacts between patients that are unrelated to bed residence. Despite the OMT’s efforts, infection and transmission events may have been missed. Ongoing serological studies performed in NSLHD and other jurisdictions may enhance our understanding.

In summary, we describe a hospital outbreak of COVID-19 linked to a community ACF cluster and which had substantial impacts on both patients and staff. Our results add weight to the recommendation that nebulisers should not be used in patients suspected to have COVID-19. Our study also illustrates that tackling viral outbreaks requires a sophisticated, detailed approach in which multidisciplinary team members must collaborate closely to achieve success. These principles will serve us well for tackling the COVID-19 pandemic in all settings.

## Supporting information

SARS-CoV-2 sequencing

## Data Availability

All data referred to in the manuscript is available.

## Acknowledgements

Our team gratefully acknowledge NSW Health Pathology-ICPMR for provision of the sequence for one of the patients. We are also grateful to the Microbiology Scientists at NSW Health Pathology, Infection Control Team at Ryde Hospital, the Ryde Executive Team and the Northern Sydney Local Health District Public Health and Executive Teams.

