## Supplementary material for "Epidemiological and Genomic analysis of a Sydney Hospital COVID-19 Outbreak": SARS-CoV-2 sequencing

### **Supporting Information**

### **SARS-CoV-2 sequencing**

RNA was extracted using the Roche MagNA Pure DNA and total NA kit on an automated extraction instrument (MagNA Pure 96). cDNA was generated with 8μl of RNA added to the Superscript IV VILO Master Mix (Thermo Fisher), which contains both random hexamers and oligo-dT primers. Amplification was then performed with the Platinum SuperFi Green PCR Mastermix (Thermo Fisher) and 1.2μl of cDNA was added to each of the 14 separate PCRs that contained published primer sets that covered the genome in a tiled approach and generated an amplicon ~2.5kb(1). All 14 amplicon products from a given sample were then pooled at equal abundance and barcoded with the ONT Rapid Barcoding Kit (SQK-RBP004) according to manufacturer’s protocol. Up to twelve samples were multiplexed on a 9.4.1 or PRO002 flow-cell and sequenced on a GridION X5 or PromethION P24 device, respectively. The *RAMPART* software package was used to monitor sequencing performance in real-time, with runs proceeding until a minimum ~200-fold coverage was achieved across all amplicons. The resulting reads were basecalled using *Guppy* (3.6) and aligned to the Wuhan Hu 1 reference genome (MN908947.3) using *minimap2* (2.17-r941). The ARTIC tool *align_trim* (<https://github.com/artic-network/artic-ncov2019>) was used to trim primer sequences from the termini of read alignments and cap sequencing depth at a maximum of 400-fold coverage. Consensus-level single nucleotide variants (SNVs) were identified using a combination of *Medaka* (0.11.5) and *Longshot*, then filtered with the ARTIC tool *artic vcf filter*(2). Consensus genome sequences were then generated by incorporating filtered SNVs into the reference genome using *bcftools consensus*.

SARS-CoV-2 lineages were determined on the full length and partial genomes with the Pangolin COVID-19 Lineage Assigner online tool (https://pangolin.cog-uk.io). Full viral genomes (n=80) were aligned with MUSCLE in Geneious Prime (<https://www.geneious.com>) and phylogenetic trees on these data were estimated using the maximum likelihood method available in the the PhyML add-on package in Geneious Prime, employing the GTR model of nucleotide substitution and 1000 bootstrap replicates. Strains representing different Pangolin lineages were obtained from GISAID and used to provide phylogenetic context. Trees were visualised with FigTree v1.4.4 and rooted on the midpoint. Strains representing different Pangolin lineages were obtained from GISAID (<https://www.gisaid.org/>)(3).
